# The Parkinson’s Disease DNA Variant Browser

**DOI:** 10.1101/2020.08.11.20172882

**Authors:** Jonggeol J. Kim, Mary B. Makarious, Sara Bandres-Ciga, J. Raphael Gibbs, Jinhui Ding, Dena G. Hernandez, Janet Brooks, Francis P. Grenn, Hirotaka Iwaki, Andrew B. Singleton, Mike A. Nalls, Cornelis Blauwendraat, on behalf of the International Parkinson’s Disease Genomics Consortium (IPDGC)

## Abstract

Parkinson’s disease (PD) is a genetically complex neurodegenerative disease with ~20 genes known to contain mutations that cause PD or atypical parkinsonism and 90 common genetic risk factors. Large-scale next-generation sequencing projects have revolutionized genomics research. Applying these data to PD, many genes have been reported to contain putative disease-causing mutations. In most instances, however, the results remain quite limited and rather preliminary, in large part because of an inability of any single group to validate findings in a large independent series of sequenced patients. We present here the Parkinson’s Disease Sequencing Browser: a Shiny-based web application that presents comprehensive summary-level frequency data from multiple large-scale genotyping and sequencing projects. The data is aggregated and involves a total of 102,127 participants, including 30,103 PD cases (including 1,650 proxy cases) and 72,024 controls. Our aim is to assist researchers on their search for PD-risk genes and variant candidates with an easily accessible and open summary-level genomic data browser for the PD research community, https://pdgenetics.shinyapps.io/VariantBrowser/.

## Introduction

Parkinson’s Disease (PD) is a neurodegenerative disease hallmarked by dopaminergic neuron degradation and Lewy-Body occlusions in the brain. The exact molecular mechanisms underlying PD remain largely unknown, but the disease is influenced by age, environmental, and complex genetic factors. Putative deleterious and highly functional variants in over 20 genes and 90 common genetic risk variants have been associated with PD or atypical parkinsonism. However, the population risk of known mutations and risk loci only represents a fraction of the known detectable heritable component of disease, suggesting that additional genetic influence is yet to be identified [1,2]. Most genes associated with PD have been discovered through linkage mapping studies in large family studies, such as *SNCA* [3] and *LRRK2* [4-6]. Some studies contain large sequencing cohort validation analyses such as the one nominating *VPS13C* [7]. The majority of the recent studies that nominate potential PD genes lack replication of results. Current research aggregation resources such as ClinVar are useful for searching known pathogenic variants, but the information presented often misses the context behind the clinical interpretation and lacks large case-control frequency information. While other resources such as MDSGene (https://www.mdsgene.org/) provide in-depth genotype-phenotype information however are lacking large study case-control frequencies [8].

Next-generation sequencing has produced petabytes of genomic data and has transformed genomic medicine. However, databases housing these data, such as gnomAD [9] and BRAVO variant browser [10], do not contain disease-specific data and there is a need for accessible resources that specifically include allele frequencies per disease group. Here, we aggregated multiple genomic datasets based on PD cases and controls and created an exonic summary data user-friendly browser: https://pdgenetics.shinyapps.io/VariantBrowser/.

## Methods

### Data Aggregation

We collected sequencing data from multiple different sources [Table 1]. The PD Genome Project includes publicly available whole-genome sequencing data from AMP-PD (https://amp-pd.org/) and other sources. The IPDGC cohort from the PDGSC data was downloaded in November 2019 and was processed using a previously described pipeline: https://github.com/ipdgc/pdgsc. The IPDGC resequencing project is a resequencing dataset that includes regions of all monogenic genes and GWAS loci regions from a previous PD GWAS [11]. The IPDGC genotype data was processed using a previously described quality control pipeline that has been previously described here: https://github.com/neurogenetics/GWAS-pipeline [1,12]. It was imputed using the Haplotype Reference Consortium Panel and filtered with the R^2^ threshold of 0.8. UK Biobank (UKB) exome data (field 23160 “Population-level FE variants, PLINK format”) was downloaded in May 2019 [13]. The PD status of the UKB participants was based on UKB field number 42033 “Source of parkinson’s disease report”, which determined the PD status on three criterias: self-report, hospital admission, and death registries. UKB proxy cases were defined as participants with no PD but with a parent with PD based on UKB field numbers 20107 and 20110 “Illnesses of father” and “Illnesses of mother”. Additional quality control was done to remove participants without case-control status and mean depth of less than 20. Note that the vast majority of data is from European ancestry. Data were trimmed to exome calling regions identical to those used in gnomAD (link), specifically bait-covered regions plus 50 bp upstream and downstream. Before merging, all hg38 data were mapped to hg19 using CrossMap v0.4.0 [14] and duplicate samples were removed based on either sample ID or PIHAT values >0.8 using PLINK v1.9 [15] (Supplementary Figure 1). The data were merged and allele count and frequencies were generated using PLINK. Merged data were annotated using ANNOVAR [16] [Figure 1].

**Table 1:** Description of the data cohorts. WGS = whole genome sequencing, WES = whole exome sequencing. * this includes AMP-PD genome data (https://amp-pd.org/)

| Study | Data Type | Cases (N) | Controls (N) | Total (N) |
| --- | --- | --- | --- | --- |
| PD Genome Project* | WGS | 2,745 | 4,071 | 6,816 |
| IPDGC Exomes | WES | 2,110 | 2,978 | 5,088 |
| IPDGC Resequencing project | Resequencing data | 2,072 | 2,818 | 4,890 |
| IPDGC GWAS Cohort | Imputed array data | 21,412 | 23,894 | 45,306 |
| UK Biobank | WES | 114 | 38,263 | 40,027 |
| *UK Biobank (proxy cases)* | WES | 1,650 | *NA* | *NA* |
| **TOTAL** |  | **30,103** | **72,024** | **102,127** |

**Figure 1:**
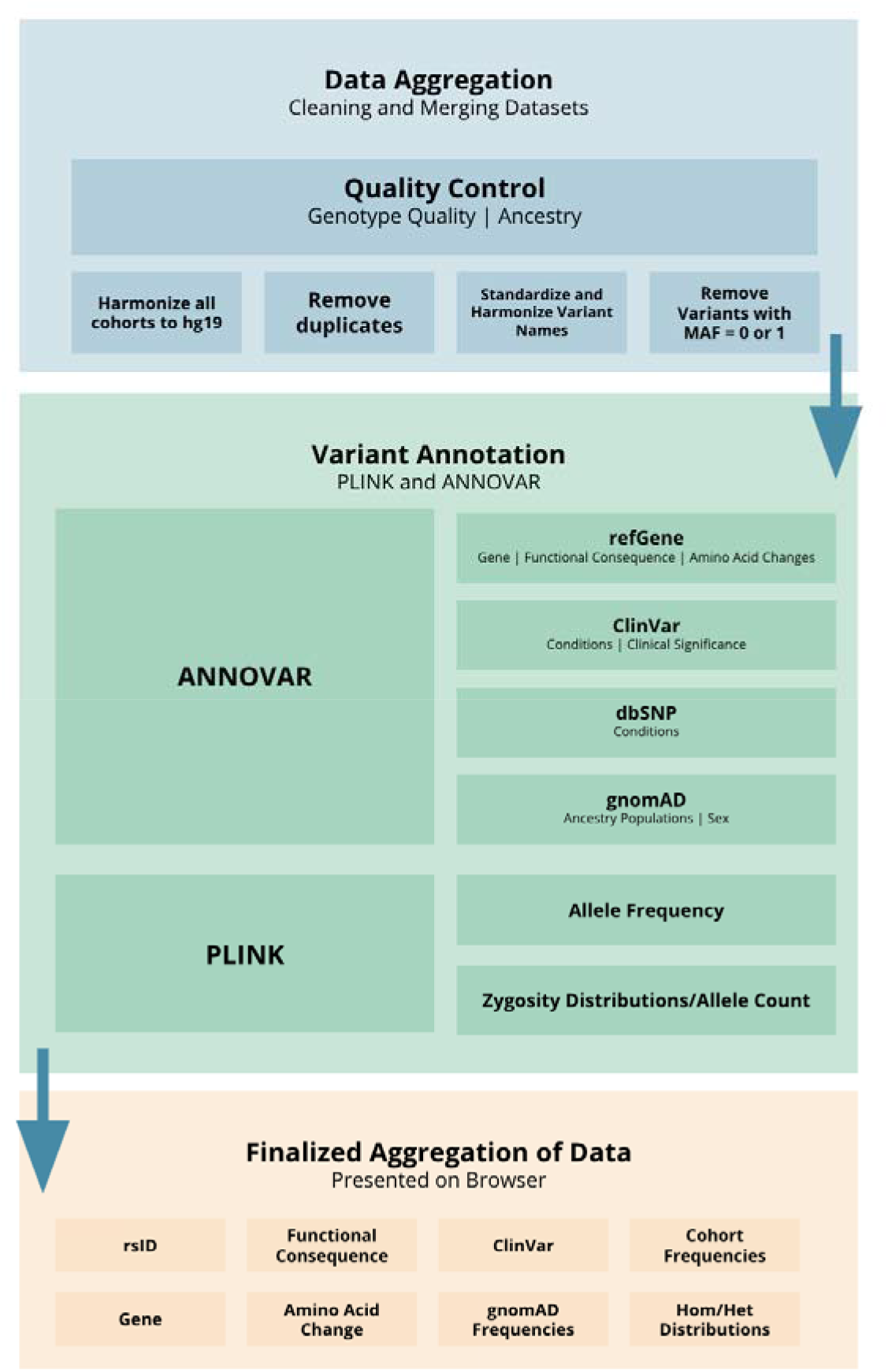
Parkinson’s Disease Sequencing Browser processing pipeline flowchart.

### Browser Design

The IPDGC Sequencing Browser was designed using the Shiny library under R version 3.6.1. All data present in the browser are non-identifiable aggregate summary-level data. The design was inspired by the gnomAD and BRAVO variant browsers, featuring gene-level information panel and separate variant level windows. However, this browser increases the information density presented in a single page format with collapsible and information panels to search results facilitating the user visualization and interpretation. It also contains an integrated tutorial function to guide new users. The browser is an open source project and the code is available on our GitHub platform: https://github.com/kimjonggeolj/ipdgc_exome_browser.

## Results

After quality control, we included a total of 6,600,433 variants from 102,127 participants, specifically 30,103 PD cases, (1,650 of which were proxy cases from UKB), and 72,024 controls. Out of the 3,581,752 exonic variants (~58%), 2,144,252 (~60%) variants were nonsynonymous variants and 1,078,618 (~30%) were synonymous variants (Supplementary Table 1). As a positive control, we assessed the allele frequency of *LRRK2* p.G2019S (rs34637584). This variant is one of the most common PD genetic factors associated with both familial and sporadic forms of disease [17]. Our browser shows the minor allele frequency of this variant at 0.007319 for cases, 0.0005717 for controls, and 0.001212 for proxy cases. Zygosity distributions show a similar pattern. Out of 22,067 cases and 63,644 controls, there are 321 heterozygous cases and 74 heterozygous controls. There is one homozygous carrier case while there are no controls with the same zygosity pattern. An association test (chi-square allelic test) without adjusting by any covariate but excluding proxy cases shows an odds ratio of 12.89 (95% CI 10.01-16.6) and a p-value of 8.57E^-145^, which is in line with previous reports for this variant. The result shows confidence that the dataset can be used to identify or provide evidence for a potential PD causal variant.

## Discussion

Here we present the Parkinson’s Disease DNA Variant browser, a public platform for the scientific community that allows rapid querying of specific genes and variants in several large case-control cohorts. Providing with a gene name or gene boundaries, the browser will present the user with summarized information on the variants found within the gene, such as the distribution of variants categorized by their functional consequences. Given a specific variant, the browser will present the user with annotated information on the variant including allele frequency, ClinVar information, and functional consequence. These functionalities can be used for example when assessing the frequency of a variant of interest identified in a PD case or within a family. As shown in the results of the *LRRK2* p.G2019S positive control example, allele frequency and zygosity distribution can give a researcher an idea of whether or not the reported risk variant may be enriched in PD cases.

Although we performed extensive quality control to ensure high-quality information was used, the data presented in the browser has inherent limitations. The datasets merged use different sequencing technologies including whole genome sequencing, whole-exome sequencing, resequencing, and imputed array-based genotyping, and that were aligned using different genome builds such as hg19 and hg38. This leaves gaps from low-imputation regions and cross-mapping failures, although the cross-mapping introduced less than 0.1% reduction in the total number of variants (Supplementary Figure 1). Furthermore, the data presented only include exonic regions and their immediate flank, thus it cannot provide information on the majority of the noncoding variants. Additionally, we only included a very limited amount of phenotypic data with case-control status which creates a potential bias for age-related penetrance. In future versions, we aim to include age of onset and other phenotype data if available. Of note, the majority of the data included is from European ancestry. We hope in future versions to increase the diversity of the data. Lastly, some genes, regions of interest, and structural variation are very complicated to genotype and sequence (including the *GBA1* gene due to the high similarities with the pseudogene) and therefore interpretation of these complex regions should be done with caution. Future larger scale and targeted studies will hopefully resolve the issue with complex genomic regions.

In summary, we present here an online resource developed for the PD research community to quickly retrieve annotated genomic information on genes and variants in a user-friendly manner, without any required data science or coding experience. Users can access the browser to get information on reported PD risk factors or supplement their own research with data from a large-scale dataset. We envisage this browser to be the first step towards easy sharing genomic information that will be continuously updated as new data becomes available.

## Data Availability

All aggregate data is freely available for use through the browser: https://pdgenetics.shinyapps.io/VariantBrowser/
The code for the browser is open source and available on the github repository here: https://github.com/kimjonggeolj/ipdgc_exome_browser/

https://pdgenetics.shinyapps.io/VariantBrowser/

## Supplementary Table and Figure

**Supplementary Table 1:** Summary of functional consequence of across variants. *Only variants with condition “Parkinson Disease” and interpretation “Pathogenic”

|  | All Variants |  | Exonic Variants |  | ClinVar Pathogenic Variants* |  |
| --- | --- | --- | --- | --- | --- | --- |
| Functional Consequence | Counts | Percentage | Counts | Percentage | Counts | Percentage |
| NA | 2,584,485 | 42.18% | 39,550 | 1.10% | 0 | 0.00% |
| Unknown | 91,916 | 1.50% | 91,916 | 2.57% | 0 | 0.00% |
| Synonymous SNV | 1,078,618 | 17.61% | 1,078,618 | 30.11% | 1 | 3.03% |
| Nonsynonymous SNV | 2,144,252 | 35.00% | 2,144,252 | 59.87% | 24 | 72.73% |
| Stopgain | 69,777 | 1.14% | 69,777 | 1.95% | 3 | 9.09% |
| Stoploss | 3168 | 0.05% | 3168 | 0.09% | 0 | 0.00% |
| Frameshift insertion | 33,643 | 0.55% | 33,643 | 0.94% | 2 | 6.06% |
| Frameshift deletion | 64,170 | 1.05% | 64,170 | 1.79% | 3 | 9.09% |
| Frameshift substitution | 88 | 0.00% | 88 | 0.00% | 0 | 0.00% |
| Nonframeshift insertion | 19,605 | 0.32% | 19,605 | 0.55% | 0 | 0.00% |
| Nonframeshift deletion | 36,934 | 0.60% | 36,934 | 1.03% | 0 | 0.00% |
| Nonframeshift substitution | 31 | 0.00% | 31 | 0.00% | 0 | 0.00% |
| Total | 6,126,687 | 100.00% | 3,581,752 | 100.00% | 33 | 100.00% |

**Supplementary Figure 1:**
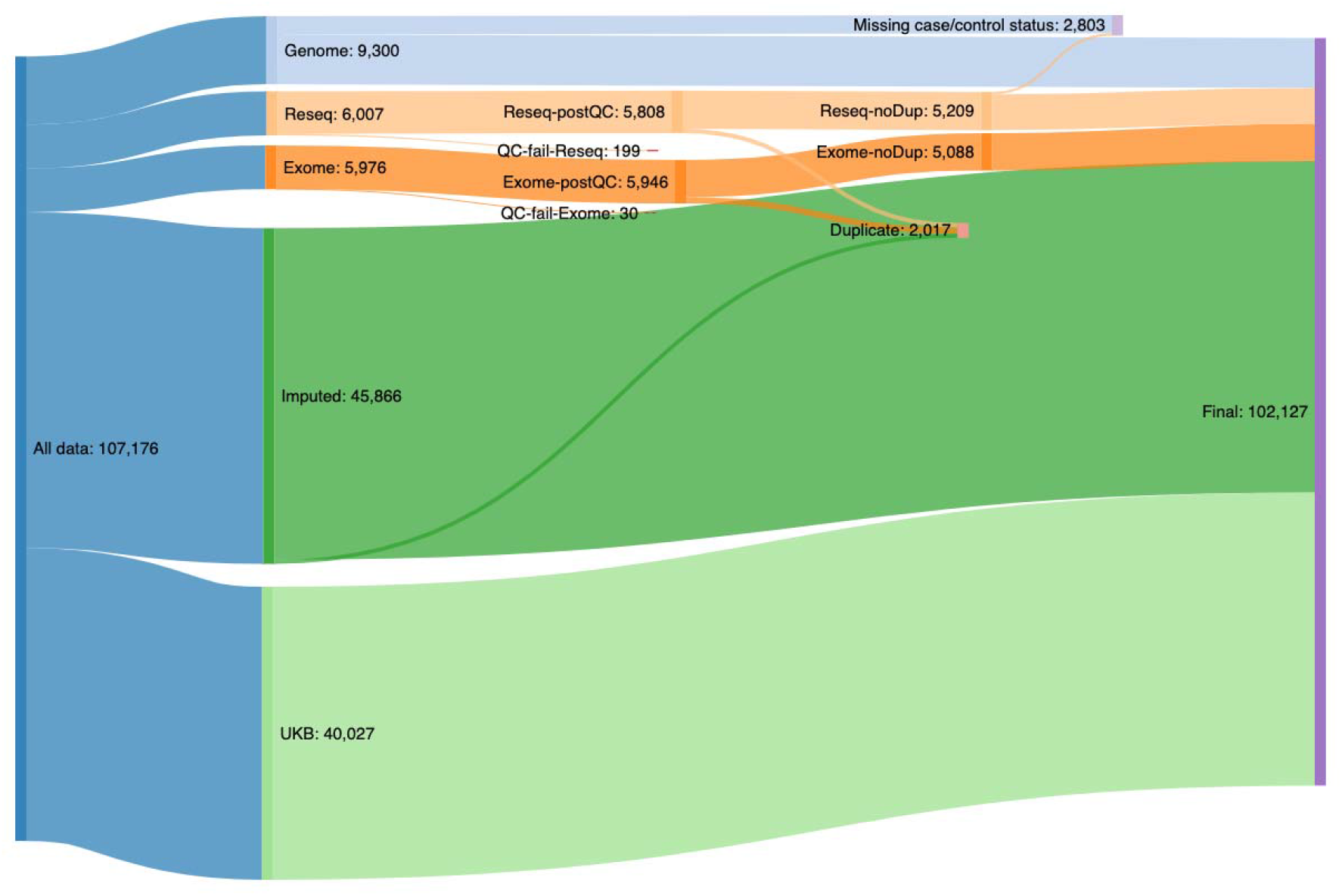
Sankey diagram of the participant filtering pipeline

## Acknowledgements and Funding

We would like to thank all of the subjects who donated their time and biological samples to be part of this study. The authors would also like to thank the Genome Aggregation Database (gnomAD) and the groups that provided exome and genome variant data to this resource. A full list of contributing groups can be found at https://gnomad.broadinstitute.org/about. Data used in the preparation of this article were obtained from the AMP PD Knowledge Platform. For up-to-date information on the study, https://www.amp-pd.org. AMP PD - a public-private partnership - is managed by the FNIH and funded by Celgene, GSK, the Michael J. Fox Foundation for Parkinson’s Research, the National Institute of Neurological Disorders and Stroke, Pfizer, Sanofi, and Verily. Clinical data used in preparation of this article were obtained from the Fox Investigation for New Discovery of Biomarkers (BioFIND), the Harvard Biomarker Study (HBS), the Parkinson’s Progression Markers Initiative (PPMI), and the Parkinson’s Disease Biomarkers Program (PDBP). BioFIND is sponsored by The Michael J. Fox Foundation for Parkinson’s Research (MJFF) with support from the National Institute for Neurological Disorders and Stroke (NINDS). The BioFIND Investigators have not participated in reviewing the data analysis or content of the manuscript. For up-to-date information on the study, visit michaeljfox.org/biofind. The Harvard NeuroDiscovery Biomarker Study (HBS) is a collaboration of HBS investigators. A full list of HBS investigator can be found at https://www.bwhparkinsoncenter.org/biobank/ and funded through philanthropy and NIH and Non-NIH funding sources. The HBS Investigators have not participated in reviewing the data analysis or content of the manuscript. PPMI - a public-private partnership - is funded by the Michael J. Fox Foundation for Parkinson’s Research and funding partners, including AbbVie, Allergan, Amathus Therapeutics, Avid Radiopharmaceuticals, Biogen, BioLegend, Bristol-Myers Squibb, Calico, Celgene, Denali, GE Healthcare, Genentech, GlaxoSmithKline, Golub Capital, Handling’s Therapeutics, Insitro, Janssen Pharmaceutica, Eli Lilly and Company, Lundbeck, Merck, Meso Scale Discovery, Neurocrine Biosciences, Pfizer, Piramal Imaging, Prevail Therapeutics, Roche, Sanofi Genzyme, Servier, Takeda Pharmaceutical Company, Teva, UCB, Verily, and Voyager Therapeutics. The PPMI Investigators have not participated in reviewing the data analysis or content of the manuscript. For up-to-date information on the study, visit www.ppmi-info.org. Parkinson’s Disease Biomarker Program (PDBP) consortium is supported by the National Institute of Neurological Disorders and Stroke (NINDS) at the National Institutes of Health. A full list of PDBP investigators can be found at https://pdbp.ninds.nih.gov/policy. The PDBP Investigators have not participated in reviewing the data analysis or content of the manuscript.. This work was supported by the Intramural Research Programs of the National Institute on Aging (NIA). This research has been conducted using the UK Biobank Resource under Application Number 33601. This study used the high-performance computational capabilities of the Biowulf Linux cluster at the National Institutes of Health (http://hpc.nih.gov). The authors also would like to thank all members of the International Parkinson Disease Genomics Consortium (IPDGC). See for a complete overview of members, acknowledgments and funding http://pdgenetics.org/partners.

## Full IPDGC author list

**United Kingdom:** Alastair J Noyce (Preventive Neurology Unit, Wolfson Institute of Preventive Medicine, QMUL, London, UK and Department of Molecular Neuroscience, UCL, London, UK), Rauan Kaiyrzhanov (Department of Molecular Neuroscience, UCL Institute of Neurology, London, UK), Ben Middlehurst (Institute of Translational Medicine, University of Liverpool, Liverpool, UK), Demis A Kia (UCL Genetics Institute; and Department of Molecular Neuroscience, UCL Institute of Neurology, London, UK), Manuela Tan (Department of Clinical Neuroscience, University College London, London, UK), Henry Houlden (Department of Molecular Neuroscience, UCL Institute of Neurology, London, UK), Catherine Storm (Department of Clinical and Movement Neurosciences, UCL Queen Square Institute of Neurology, London, UK), Huw R Morris (Department of Clinical Neuroscience, University College London, London, UK), Helene Plun-Favreau (Department of Molecular Neuroscience, UCL Institute of Neurology, London, UK), Peter Holmans (Biostatistics & Bioinformatics Unit, Institute of Psychological Medicine and Clinical Neuroscience, MRC Centre for Neuropsychiatric Genetics & Genomics, Cardiff, UK), John Hardy (Department of Molecular Neuroscience, UCL Institute of Neurology, London, UK), Daniah Trabzuni (Department of Molecular Neuroscience, UCL Institute of Neurology, London, UK; Department of Genetics, King Faisal Specialist Hospital and Research Centre, Riyadh, 11211 Saudi Arabia), John Quinn (Institute of Translational Medicine, University of Liverpool, Liverpool, UK), Vivien Bubb (Institute of Translational Medicine, University of Liverpool, Liverpool, UK), Kin Y Mok (Department of Molecular Neuroscience, UCL Institute of Neurology, London, UK), Kerri J. Kinghorn (Institute of Healthy Ageing, Research Department of Genetics, Evolution and Environment, University College London, London, UK), Nicholas W Wood (UCL Genetics Institute; and Department of Molecular Neuroscience, UCL Institute of Neurology, London, UK), Patrick Lewis (University of Reading, Reading, UK), Sebastian R Schreglmann (Department of Molecular Neuroscience, UCL Institute of Neurology, London, UK), Ruth Lovering (University College London, London, UK), Lea R’Bibo (Department of Molecular Neuroscience, UCL Institute of Neurology, London, UK), Claudia Manzoni (University of Reading, Reading, UK), Mie Rizig (Department of Molecular Neuroscience, UCL Institute of Neurology, London, UK), Mina Ryten (Department of Molecular Neuroscience, UCL Institute of Neurology, London, UK), Sebastian Guelfi (Department of Molecular Neuroscience, UCL Institute of Neurology, London, UK), Valentina Escott-Price (MRC Centre for Neuropsychiatric Genetics and Genomics, Cardiff University School of Medicine, Cardiff, UK), Viorica Chelban (Department of Molecular Neuroscience, UCL Institute of Neurology, London, UK), Thomas Foltynie (UCL Institute of Neurology, London, UK), Nigel Williams (MRC Centre for Neuropsychiatric Genetics and Genomics, Cardiff, UK), Karen E. Morrison (Faculty of Medicine, University of Southampton, UK), Carl Clarke (University of Birmingham, Birmingham, UK and Sandwell and West Birmingham Hospitals NHS Trust, Birmingham, UK), Kirsten Harvey (UCL School of Pharmacy, UK), Benjamin M Jacobs (Preventive Neurology Unit, Wolfson Institute of Preventive Medicine, QMUL, London, UK).

**France:** Alexis Brice (Institut du Cerveau et de la Moelle épinière, ICM, Inserm U 1127, CNRS, UMR 7225, Sorbonne Universités, UPMC University Paris 06, UMR S 1127, AP-HP, Pitié-Salpêtrière Hospital, Paris, France), Fabrice Danjou (Institut du Cerveau et de la Moelle épinière, ICM, Inserm U 1127, CNRS, UMR 7225, Sorbonne Universités, UPMC University Paris 06, UMR S 1127, AP-HP, Pitié-Salpêtrière Hospital, Paris, France), Suzanne Lesage (Institut du Cerveau et de la Moelle épinière, ICM, Inserm U 1127, CNRS, UMR 7225, Sorbonne Universités, UPMC University Paris 06, UMR S 1127, AP-HP, Pitié-Salpêtrière Hospital, Paris, France), Jean-Christophe Corvol (Institut du Cerveau et de la Moelle épinière, ICM, Inserm U 1127, CNRS, UMR 7225, Sorbonne Universités, UPMC University Paris 06, UMR S 1127, Centre d’Investigation Clinique Pitié Neurosciences CIC-1422, AP-HP, Pitié-Salpêtrière Hospital, Paris, France), Maria Martinez (INSERM UMR 1220; and Paul Sabatier University, Toulouse, France),

**Germany:** Claudia Schulte (Department for Neurodegenerative Diseases, Hertie Institute for Clinical Brain Research, University of Tübingen, and DZNE, German Center for Neurodegenerative Diseases, Tübingen, Germany), Kathrin Brockmann (Department for Neurodegenerative Diseases, Hertie Institute for Clinical Brain Research, University of Tübingen, and DZNE, German Center for Neurodegenerative Diseases, Tübingen, Germany), Javier Simón-Sánchez (Department for Neurodegenerative Diseases, Hertie Institute for Clinical Brain Research, University of Tübingen, and DZNE, German Center for Neurodegenerative Diseases, Tübingen, Germany), Peter Heutink (DZNE, German Center for Neurodegenerative Diseases and Department for Neurodegenerative Diseases, Hertie Institute for Clinical Brain Research, University of Tübingen, Tübingen, Germany), Patrizia Rizzu (DZNE, German Center for Neurodegenerative Diseases), Manu Sharma (Centre for Genetic Epidemiology, Institute for Clinical Epidemiology and Applied Biometry, University of Tubingen, Germany), Thomas Gasser (Department for Neurodegenerative Diseases, Hertie Institute for Clinical Brain Research, and DZNE, German Center for Neurodegenerative Diseases, Tübingen, Germany), Susanne A. Schneider (Department of Neurology, Ludwig-Maximilians-University Munich, München, Germany)

**United States of America:** Mark R Cookson (Laboratory of Neurogenetics, National Institute on Aging, Bethesda, USA), Sara Bandres-Ciga (Laboratory of Neurogenetics, National Institute on Aging, Bethesda, MD, USA), Cornelis Blauwendraat (Laboratory of Neurogenetics, National Institute on Aging, Bethesda, MD, USA), David W. Craig (Department of Translational Genomics, Keck School of Medicine, University of Southern California, Los Angeles, USA), Kimberley Billingsley (Laboratory of Neurogenetics, National Institute on Aging, Bethesda, MD, USA), Mary B. Makarious (Laboratory of Neurogenetics, National Institute on Aging, Bethesda, MD, USA), Derek P. Narendra (Inherited Movement Disorders Unit, National pInstitute of Neurological Disorders and Stroke, Bethesda, MD, USA), Faraz Faghri (Laboratory of Neurogenetics, National Institute on Aging, Bethesda, USA; Department of Computer Science, University of Illinois at Urbana-Champaign, Urbana, IL, USA), J Raphael Gibbs (Laboratory of Neurogenetics, National Institute on Aging, National Institutes of Health, Bethesda, MD, USA), Dena G. Hernandez (Laboratory of Neurogenetics, National Institute on Aging, Bethesda, MD, USA), Kendall Van Keuren-Jensen (Neurogenomics Division, TGen, Phoenix, AZ USA), Joshua M. Shulman (Departments of Neurology, Neuroscience, and Molecular & Human Genetics, Baylor College of Medicine, Houston, Texas, USA; Jan and Dan Duncan Neurological Research Institute, Texas Children’s Hospital, Houston, Texas, USA), Hirotaka Iwaki (Laboratory of Neurogenetics, National Institute on Aging, Bethesda, MD, USA), Hampton L. Leonard (Laboratory of Neurogenetics, National Institute on Aging, Bethesda, MD, USA), Mike A. Nalls (Laboratory of Neurogenetics, National Institute on Aging, Bethesda, USA; CEO/Consultant Data Tecnica International, Glen Echo, MD, USA), Laurie Robak (Baylor College of Medicine, Houston, Texas, USA), Jose Bras (Center for Neurodegenerative Science, Van Andel Research Institute, Grand Rapids, Michigan, USA), Rita Guerreiro (Center for Neurodegenerative Science, Van Andel Research Institute, Grand Rapids, Michigan, USA), Steven Lubbe (Ken and Ruth Davee Department of Neurology and Simpson Querrey Center for Neurogenetics, Northwestern University Feinberg School of Medicine, Chicago, IL, USA), Steven Finkbeiner (Departments of Neurology and Physiology, University of California, San Francisco; Gladstone Institute of Neurological Disease; Taube/Koret Center for Neurodegenerative Disease Research, San Francisco, CA, USA), Niccolo E. Mencacci (Northwestern University Feinberg School of Medicine, Chicago, IL, USA), Codrin Lungu (National Institutes of Health Division of Clinical Research, NINDS, National Institutes of Health, Bethesda, MD, USA), Andrew B Singleton (Laboratory of Neurogenetics, National Institute on Aging, Bethesda, MD, USA), Sonja W. Scholz (Neurodegenerative Diseases Research Unit, National Institute of Neurological Disorders and Stroke, Bethesda, MD, USA), Xylena Reed (Laboratory of Neurogenetics, National Institute on Aging, Bethesda, MD, USA). Roy N. Alcalay (Department of Neurology, College of Physicians and Surgeons, Columbia University Medical Center, New York, NY, USA, Taub Institute for Research on Alzheimer’s Disease and the Aging Brain, College of Physicians and Surgeons, Columbia University Medical Center, New York, NY, USA). Zbigniew K. Wszolek (Department of Neurology, Mayo Clinic Jacksonville, FL, USA), Ryan J. Uitti (Department of Neurology, Mayo Clinic Jacksonville, FL, USA), Owen A. Ross (Departments of Neuroscience & Clinical Genomics, Mayo Clinic Jacksonville, FL, USA), Francis P. Grenn (Laboratory of Neurogenetics, National Institute on Aging, Bethesda, MD, USA).

**Canada:** Ziv Gan-Or (Montreal Neurological Institute and Hospital, Department of Neurology & Neurosurgery, Department of Human Genetics, McGill University, Montréal, QC, H3A 0G4, Canada), Guy A. Rouleau (Montreal Neurological Institute and Hospital, Department of Neurology & Neurosurgery, Department of Human Genetics, McGill University, Montréal, QC, H3A 0G4, Canada), Lynne Krohn (Montreal Neurological Institute and Hospital, Department of Neurology & Neurosurgery, Department of Human Genetics, McGill University, Montréal, QC, H3A 0G4, Canada), Kheireddin Mufti (Montreal Neurological Institute and Hospital, Department of Neurology & Neurosurgery, Department of Human Genetics, McGill University, Montréal, QC, H3A 0G4, Canada),

**The Netherlands:** Jacobus J van Hilten (Department of Neurology, Leiden University Medical Center, Leiden, Netherlands), Johan Marinus (Department of Neurology, Leiden University Medical Center, Leiden, Netherlands)

**Spain:** Astrid D. Adarmes-Gómez (Instituto de Biomedicina de Sevilla (IBiS), Hospital Universitario Virgen del Rocío/CSIC/Universidad de Sevilla, Seville), Miquel Aguilar (Fundació Docència i Recerca Mútua de Terrassa and Movement Disorders Unit, Department of Neurology, University Hospital Mutua de Terrassa, Terrassa, Barcelona.), Ignacio Alvarez (Fundació Docència i Recerca Mútua de Terrassa and Movement Disorders Unit, Department of Neurology, University Hospital Mutua de Terrassa, Terrassa, Barcelona.),Victoria Alvarez (Hospital Universitario Central de Asturias, Oviedo), Francisco Javier Barrero (Hospital Universitario San Cecilio de Granada, Universidad de Granada), Jesùs Alberto Bergareche Yarza (Instituto de Investigación Sanitaria Biodonostia, San Sebastián), Inmaculada Bernal-Bernal (Instituto de Biomedicina de Sevilla (IBiS), Hospital Universitario Virgen del Rocío/CSIC/Universidad de Sevilla, Seville), Marta Blazquez (Hospital Universitario Central de Asturias, Oviedo), Marta Bonilla-Toribio (Instituto de Biomedicina de Sevilla (IBiS), Hospital Universitario Virgen del Rocío/CSIC/Universidad de Sevilla, Seville), Juan A. Botía (Universidad de Murcia, Murcia), María Teresa Boungiorno (Fundació Docència i Recerca Mùtua de Terrassa and Movement Disorders Unit, Department of Neurology, University Hospital Mutua de Terrassa, Terrassa, Barcelona.) Dolores Buiza-Rueda (Instituto de Biomedicina de Sevilla (IBiS), Hospital Universitario Virgen del Rocío/CSIC/Universidad de Sevilla, Seville), Ana Cámara (Hospital Clinic de Barcelona), Fátima Carrillo (Instituto de Biomedicina de Sevilla (IBiS), Hospital Universitario Virgen del Rocío/CSIC/Universidad de Sevilla, Seville), Mario Carrión-Claro (Instituto de Biomedicina de Sevilla (IBiS), Hospital Universitario Virgen del Rocío/CSIC/Universidad de Sevilla, Seville), Debora Cerdan (Hospital General de Segovia, Segovia), Jordi Clarimón (Memory Unit, Department of Neurology, IIB Sant Pau, Hospital de la Santa Creu i Sant Pau, Universitat Autònoma de Barcelona and Centro de Investigación Biomédica en Red en Enfermedades Neurodegenerativas (CIBERNED), Madrid),Yaroslau Compta (Hospital Clinic de Barcelona), Monica Diez-Fairen (Fundació Docència i Recerca Mùtua de Terrassa and Movement Disorders Unit, Department of Neurology, University Hospital Mutua de Terrassa, Terrassa, Barcelona.), Oriol Dols-Icardo (Memory Unit, Department of Neurology, IIB Sant Pau, Hospital de la Santa Creu i Sant Pau, Universitat Autònoma de Barcelona, Barcelona, and Centro de Investigación Biomédica en Red en Enfermedades Neurodegenerativas (CIBERNED), Madrid), Jacinto Duarte (Hospital General de Segovia, Segovia), Raquel Duran (Centro de Investigacion Biomedica, Universidad de Granada, Granada), Francisco Escamilla-Sevilla (Hospital Universitario Virgen de las Nieves, Instituto de Investigación Biosanitaria de Granada, Granada), Mario Ezquerra (Hospital Clinic de Barcelona), Cici Feliz (Departmento de Neurologia, Instituto de Investigación Sanitaria Fundación Jiménez Díaz, Madrid, Spain), Manel Fernández (Hospital Clinic de Barcelona), Rubén Fernández-Santiago (Hospital Clinic de Barcelona), Ciara Garcia (Hospital Universitario Central de Asturias, Oviedo), Pedro García-Ruiz (Instituto de Investigación Sanitaria Fundación Jiménez Díaz, Madrid), Pilar Gómez-Garre (Instituto de Biomedicina de Sevilla (IBiS), Hospital Universitario Virgen del Rocío/CSIC/Universidad de Sevilla, Seville), Maria Jose Gomez Heredia (Hospital Universitario Virgen de la Victoria, Malaga), Isabel Gonzalez-Aramburu (Hospital Universitario Marqués de Valdecilla-IDIVAL, Santander), Ana Gorostidi Pagola (Instituto de Investigación Sanitaria Biodonostia, San Sebastián), Janet Hoenicka (Institut de Recerca Sant Joan de Déu, Barcelona), Jon Infante (Hospital Universitario Marqués de Valdecilla-IDIVAL and University of Cantabria, Santander, and Centro de Investigación Biomédica en Red en Enfermedades Neurodegenerativas (CIBERNED), Silvia Jesús (Instituto de Biomedicina de Sevilla (IBiS), Hospital Universitario Virgen del Rocío/CSIC/Universidad de Sevilla, Seville), Adriano Jimenez-Escrig (Hospital Universitario Ramón y Cajal, Madrid), Jaime Kulisevsky (Movement Disorders Unit, Department of Neurology, IIB Sant Pau, Hospital de la Santa Creu i Sant Pau, Universitat Autònoma de Barcelona, Barcelona, and Centro de Investigación Biomédica en Red en Enfermedades Neurodegenerativas (CIBERNED)), Miguel A. Labrador-Espinosa (Instituto de Biomedicina de Sevilla (IBiS), Hospital Universitario Virgen del Rocío/CSIC/Universidad de Sevilla, Seville), Jose Luis Lopez-Sendon (Hospital Universitario Ramón y Cajal, Madrid), Adolfo López de Munain Arregui (Instituto de Investigación Sanitaria Biodonostia, San Sebastián), Daniel Macias (Instituto de Biomedicina de Sevilla (IBiS), Hospital Universitario Virgen del Rocío/CSIC/Universidad de Sevilla, Seville), Irene Martínez Torres (Department of Neurology, Instituto de Investigación Sanitaria La Fe, Hospital Universitario y Politécnico La Fe, Valencia), Juan Marín (Movement Disorders Unit, Department of Neurology, IIB Sant Pau, Hospital de la Santa Creu i Sant Pau, Universitat Autónoma de Barcelona, Barcelona, and Centro de Investigación Biomédica en Red en Enfermedades Neurodegenerativas (CIBERNED)), Maria Jose Marti (Hospital Clinic Barcelona), Juan Carlos Martínez-Castrillo (Instituto Ramón y Cajal de Investigación Sanitaria, Hospital Universitario Ramón y Cajal, Madrid), Carlota Méndez-del-Barrio (Instituto de Biomedicina de Sevilla (IBiS), Hospital Universitario Virgen del Rocío/CSIC/Universidad de Sevilla, Seville), Manuel Menéndez González (Hospital Universitario Central de Asturias, Oviedo), Marina Mata (Department of Neurology, Hospital Universitario Infanta Sofía, Madrid, Spain) Adolfo Mínguez (Hospital Universitario Virgen de las Nieves, Granada, Instituto de Investigación Biosanitaria de Granada), Pablo Mir (Instituto de Biomedicina de Sevilla (IBiS), Hospital Universitario Virgen del Rocío/CSIC/Universidad de Sevilla, Seville), Elisabet Mondragon Rezola (Instituto de Investigación Sanitaria Biodonostia, San Sebastián), Esteban Muñoz (Hospital Clinic Barcelona), Javier Pagonabarraga (Movement Disorders Unit, Department of Neurology, IIB Sant Pau, Hospital de la Santa Creu i Sant Pau, Universitat Autónoma de Barcelona, Barcelona, and Centro de Investigación Biomédica en Red en Enfermedades Neurodegenerativas (CIBERNED)), Pau Pastor (Fundació Docència i Recerca Mútua de Terrassa and Movement Disorders Unit, Department of Neurology, University Hospital Mutua de Terrassa, Terrassa, Barcelona.), Francisco Perez Errazquin (Hospital Universitario Virgen de la Victoria, Malaga), Teresa Periñán-Tocino (Instituto de Biomedicina de Sevilla (IBiS), Hospital Universitario Virgen del Rocío/CSIC/Universidad de Sevilla, Seville), Javier Ruiz-Martínez (Hospital Universitario Donostia, Instituto de Investigación Sanitaria Biodonostia, San Sebastián), Clara Ruz (Centro de Investigacion Biomedica, Universidad de Granada, Granada), Antonio Sanchez Rodriguez (Hospital Universitario Marqués de Valdecilla-IDIVAL, Santander), María Sierra (Hospital Universitario Marqués de Valdecilla-IDIVAL, Santander), Esther Suarez-Sanmartin (Hospital Universitario Central de Asturias, Oviedo), Cesar Tabernero (Hospital General de Segovia, Segovia), Juan Pablo Tartari (Fundació Docéncia i Recerca Mútua de Terrassa and Movement Disorders Unit, Department of Neurology, University Hospital Mutua de Terrassa, Terrassa, Barcelona), Cristina Tejera-Parrado (Instituto de Biomedicina de Sevilla (IBiS), Hospital Universitario Virgen del Rocío/CSIC/Universidad de Sevilla, Seville), Eduard Tolosa (Hospital Clinic Barcelona), Francesc Valldeoriola (Hospital Clinic Barcelona), Laura Vargas-González (Instituto de Biomedicina de Sevilla (IBiS), Hospital Universitario Virgen del Rocío/CSIC/Universidad de Sevilla, Seville), Lydia Vela (Department of Neurology, Hospital Universitario Fundación Alcorcón, Madrid), Francisco Vives (Centro de Investigacion Biomedica, Universidad de Granada, Granada).

**Austria:** Alexander Zimprich (Department of Neurology, Medical University of Vienna, Austria)

**Norway:** Lasse Pihlstrom (Department of Neurology, Oslo University Hospital, Oslo, Norway), Mathias Toft (Department of Neurology and Institute of Clinical Medicine, Oslo University Hospital, Oslo, Norway)

**Estonia:** Pille Taba (Department of Neurology and Neurosurgery, University of Tartu, Tartu, Estonia)

**Australia:** Sulev Koks (Centre for Molecular Medicine and Innovative Therapeutics, Murdoch University, Murdoch, 6150, Perth, Western Australia; Perron Institute for Neurological and Translational Science, Nedlands, 6009, Perth, Western Australia)

**Israel:** Sharon Hassin-Baer (The Movement Disorders Institute, Department of Neurology and Sagol Neuroscience Center, Chaim Sheba Medical Center, Tel-Hashomer, 5262101, Ramat Gan, Israel, Sackler Faculty of Medicine, Tel Aviv University, Tel Aviv, Israel)

**Finland:** Kari Majamaa (Institute of Clinical Medicine, Department of Neurology, University of Oulu, Oulu, Finland; Department of Neurology and Medical Research Center, Oulu University Hospital, Oulu, Finland), Ari Siitonen (Institute of Clinical Medicine, Department of Neurology, University of Oulu, Oulu, Finland; Department of Neurology and Medical Research Center, Oulu University Hospital, Oulu, Finland), Pentti Tienari (Clinical Neurosciences, Neurology, University of Helsinki, Helsinki, Finland, Helsinki University Hospital, Helsinki, Finland)

**Nigeria:** Njideka U. Okubadejo (University of Lagos, Lagos State, Nigeria), Oluwadamilola O. Ojo (University of Lagos, Lagos State, Nigeria),

**Kazakhstan:** Rauan Kaiyrzhanov (Department of Molecular Neuroscience, UCL Institute of Neurology, London, UK), Chingiz Shashkin (Kazakh National Medical University named after Asfendiyarov, Almaty, Kazakhstan), Nazira Zharkinbekova (South Kazakhstan Medical Academy, Shymkent, Kazakhstan), Vadim Akhmetzhanov (Astana Medical University, Astana Kazakhstan), Akbota Aitkulova (National Center for Biotechnology, Astana, Kazakhstan; Al-Farabi Kazakh national university. Almaty Kazakhstan), Elena Zholdybayeva (National Center for Biotechnology, Astana, Kazakhstan), Zharkyn Zharmukhanov (National Center for Biotechnology, Astana, Kazakhstan), Gulnaz Kaishybayeva (Scientific and practical center “Institute of neurology named after Smagul Kaishibayev”, Almaty, Kazakhstan), Altynay Karimova (Scientific and practical center “Institute of neurology named after Smagul Kaishibayev”, Almaty, Kazakhstan), Talgat Khaibullin (Semey Medical University, Semey, Kazakhstan).

**Ireland:** Timothy L. Lynch (The Dublin Neurological Institute at the Mater Misericordiae University Hospital, Dublin, Ireland & School of Medicine and Medical Science, University College Dublin, Dublin, Ireland).

## Full IPDGC acknowledgment

We would like to thank all of the subjects who donated their time and biological samples to be a part of this study. We also would like to thank all members of the International Parkinson Disease Genomics Consortium (IPDGC). See for a complete overview of members, acknowledgements and funding http://pdgenetics.org/partners.This work was supported in part by the Intramural Research Programs of the National Institute of Neurological Disorders and Stroke (NINDS), the National Institute on Aging (NIA), and the National Institute of Environmental Health Sciences both part of the National Institutes of Health, Department of Health and Human Services; project numbers 1ZIA-NS003154, Z01- AG000949-02, Z01-ES101986 and UK ADC NIA P30-AG0-28383. In addition this work was supported by the Department of Defense (award W81XWH-09-2-0128), and The Michael J Fox Foundation for Parkinson’s Research. This work was supported by National Institutes of Health grants R01NS037167, R01CA141668, P50NS071674, American Parkinson Disease Association (APDA); Barnes Jewish Hospital Foundation; Greater St Louis Chapter of the APDA. The KORA (Cooperative Research in the Region of Augsburg) research platform was started and financed by the Forschungszentrum für Umwelt und Gesundheit, which is funded by the German Federal Ministry of Education, Science, Research, and Technology and by the State of Bavaria. This study was also funded by the German Federal Ministry of Education and Research (BMBF) under the funding code 031A430A, the EU Joint Programme - Neurodegenerative Diseases Research (JPND) project under the aegis of JPND -www.jpnd.eu- through Germany, BMBF, funding code 01ED1406 and iMed - the Helmholtz Initiative on Personalized Medicine. This study is funded by the German National Foundation grant (DFG SH599/6-1) (grant to M.S), Michael J Fox Foundation, and MSA Coalition, USA (to M.S). The French GWAS work was supported by the French National Agency of Research (ANR-08-MNP-012). This study was also funded by France-Parkinson Association, Fondation de France, the French program “Investissements d’avenir” funding (ANR-10-IAIHU-06) and a grant from Assistance Publique-Hôpitaux de Paris (PHRC, AOR- 08010) for the French clinical data. This study was also sponsored by the Landspitali University Hospital Research Fund (grant to SSv); Icelandic Research Council (grant to SSv); and European Community Framework Programme 7, People Programme, and IAPP on novel genetic and phenotypic markers of Parkinson’s disease and Essential Tremor (MarkMD), contract number PIAP-GA-2008-230596 MarkMD (to HP and JHu). We thank members of the North American Brain Expression Consortium (NABEC) for providing DNA samples derived from brain tissue. Brain tissue for the NABEC cohort was obtained from the Baltimore Longitudinal Study on Aging at the Johns Hopkins School of Medicine, the NICHD Brain and Tissue Bank for Developmental Disorders at the University of Maryland, the Banner Sun Health Research Institute Brain and Body Donation Program, and from the University of Kentucky Alzheimer’s Disease Center Brain Bank. Institutional research funding IUT20-46 was received of the Estonian Ministry of Education and Research (SK). The McGill study was funded by the Michael J. Fox Foundation and the Canadian Consortium on Neurodegeneration in Aging (CCNA). Mayo Clinic is an American Parkinson Disease Association (APDA) Mayo Clinic Information and Referral Center, an APDA Center for Advanced Research and the Mayo Clinic Lewy Body Dementia Association (LBDA) Research Center of Excellence. OAR is supported by the National Institutes of Health (NIH; R01 NS78086; U54 NS100693, U54 NS110435), the US Department of Defense (W81XWH- 17-1-0249), The Little Family Foundation, The Mayo Clinic Functional Genomics of LBD Program the Mayo Clinic Center for Individualized Medicine, and the Michael J. Fox Foundation. Z. K. Wszolek is supported by the NIH, Mayo Clinic Center for Regenerative Medicine, The Sol Goldman Charitable Trust, and Donald G. and Jodi P. Heeringa, the Haworth Family Professorship in Neurodegenerative Diseases fund, and by the Albertson Parkinson’s Research Foundation. This study utilized the high-performance computational capabilities of the Biowulf Linux cluster at the National Institutes of Health, Bethesda, Md. (http://biowulf.nih.gov), and DNA panels, samples, and clinical data from the National Institute of Neurological Disorders and Stroke Human Genetics Resource Center DNA and Cell Line Repository. People who contributed samples are acknowledged in descriptions of every panel on the repository website. We thank the French Parkinson’s Disease Genetics Study Group and the Drug Interaction with genes (DIGPD) study group: Y Agid, M Anheim, F Artaud, A-M Bonnet, C Bonnet, F Bourdain, J-P Brandel, C Brefel-Courbon, M Borg, A Brice, E Broussolle, F Cormier-Dequaire, J-C Corvol, P Damier, B Debilly, B Degos, P Derkinderen, A Destée, A Dürr, F Durif, A Elbaz, D Grabli, A Hartmann, S Klebe, P. Krack, J Kraemmer, S Leder, S Lesage, R Levy, E Lohmann, L Lacomblez, G Mangone, L-L Mariani, A-R Marques, M Martinez, V Mesnage, J Muellner, F Ory-Magne, F Pico, V Planté-Bordeneuve, P Pollak, O Rascol, K Tahiri, F Tison, C Tranchant, E Roze, M Tir, M Vérin, F Viallet, M Vidailhet, A You. We also thank the members of the French 3C Consortium: A Alpérovitch, C Berr, C Tzourio, and P Amouyel for allowing us to use part of the 3C cohort, and D Zelenika for support in generating the genome-wide molecular data. We thank P Tienari (Molecular Neurology Programme, Biomedicum, University of Helsinki), T Peuralinna (Department of Neurology, Helsinki University Central Hospital), L Myllykangas (Folkhalsan Institute of Genetics and Department of Pathology, University of Helsinki), and R Sulkava (Department of Public Health and General Practice Division of Geriatrics, University of Eastern Finland) for the Finnish controls (Vantaa85+ GWAS data). We used genome-wide association data generated by the Wellcome Trust Case-Control Consortium 2 (WTCCC2) from UK patients with Parkinson’s disease and UK control individuals from the 1958 Birth Cohort and National Blood Service. Genotyping of UK replication cases on ImmunoChip was part of the WTCCC2 project, which was funded by the Wellcome Trust (083948/Z/07/Z). UK population control data was made available through WTCCC1. This study was supported by the Medical Research Council and Wellcome Trust disease centre (grant WT089698/Z/09/Z to NW, JHa, and ASc). As with previous IPDGC efforts, this study makes use of data generated by the Wellcome Trust Case-Control Consortium. A full list of the investigators who contributed to the generation of the data is available from www.wtccc.org.uk. Funding for the project was provided by the Wellcome Trust under award 076113, 085475 and 090355. This study was also supported by Parkinson’s UK (grants 8047 and J-0804) and the Medical Research Council (G0700943 and G1100643). Sequencing and genotyping done in McGill University was supported by grants from the Michael J. Fox Foundation, the Canadian Consortium on Neurodegeneration in Aging (CCNA), the Canada First Research Excellence Fund (CFREF), awarded to McGill University for the Healthy Brains for Healthy Lives (HBHL) program and Parkinson’s Society Canada. We thank Jeffrey Barrett and Jason Downing (Illumina Inc) for assistance with the design of the ImmunoChip and NeuroX arrays. DNA extraction work that was done in the UK was undertaken at University College London Hospitals, University College London, who received a proportion of funding from the Department of Health’s National Institute for Health Research Biomedical Research Centres funding. This study was supported in part by the Wellcome Trust/Medical Research Council Joint Call in Neurodegeneration award (WT089698) to the Parkinson’s Disease Consortium (UKPDC), whose members are from the UCL Institute of Neurology, University of Sheffield, and the Medical Research Council Protein Phosphorylation Unit at the University of Dundee. We thank the Quebec Parkinson’s Network (http://rpq-qpn.org) and its members. This work was supported by the Medical Research Council grant MR/N026004/1. The Braineac project was supported by the MRC through the MRC Sudden Death Brain Bank Grant (MR/G0901254) to J.H. P.A.L. was supported by the MRC (grants MR/N026004/1 and MR/L010933/1) and Michael J. Fox Foundation for Parkinson’s Research. Mike A. Nalls’ participation is supported by a consulting contract between Data Tecnica International and the National Institute on Aging, NIH, Bethesda, MD, USA, as a possible conflict of interest Dr. Nalls also consults for Illumina Inc, Lysosomal Therapeutics Inc, the Michael J. Fox Foundation and Vivid Genomics among others.

